# One Year of Evidence on Mental Health Disorders in China during the COVID-19 Crisis - A Systematic Review and Meta-Analysis

**DOI:** 10.1101/2021.02.01.21250929

**Authors:** Xi Chen, Jiyao Chen, Meimei Zhang, Richard Z. Chen, Rebecca Kechen Dong, Zhe Dong, Yingying Ye, Lingyao Tong, Bryan Z. Chen, Ruiying Zhao, Wenrui Cao, Peikai Li, Stephen X. Zhang

## Abstract

This paper provides a systematic review and meta-analysis on the prevalence rate of mental health issues of general population, general and frontline healthcare workers (HCWs) in China over one year of the COVID-19 crisis. We systematically searched PubMed, Embase, Web of Science, and Medrxiv at November 16th, 2020, pooled data using random-effects meta-analyses to estimate the prevalence rates, and ran meta-regression to tease out the heterogeneity. The meta-regression results uncovered several predictors of the prevalence rates, including severity, type of mental issues, population, sampling location, and study quality. Pooled prevalence rates are significantly different from, yet largely between, the findings of previous meta-analyses, suggesting the results of our larger study are consistent with yet more accurate than the findings of the smaller, previous meta-analyses. The prevalence rates of distress and insomnia and those of frontline HCWs are higher suggest future research and interventions should pay more attention to those mental outcomes and populations. Our findings suggest a need to examine the prevalence rates at varying levels of severity. The one-year cumulative evidence on sampling locations (Wuhan vs. non-Wuhan) corroborates the typhoon eye effect theory.

**Trial registration:** CRD4202022059

## 1. INTRODUCTION

Since its first publicly known cases in Wuhan, China, on November 17, 2019, the COVID-19 (coronavirus disease 2019) crisis has become one of the worst epidemics in human record (World Health Organization, 2020). The sudden outburst of this highly infectious disease and the containment measures such as quarantine and social distancing have posed an unprecedented disruption on the life and work of the general population and healthcare workers (HCWs) (Douglas et al., 2020; Zhang et al., 2020h). Their mental health conditions under the COVID-19 epidemic have been documented first and most extensively to date in China (Bareeqa et al., 2020; Pappa, 2020). The accumulating number of such studies has triggered several rapid meta-analyses (Bareeqa et al., 2020; Kisely et al., 2020; Krishnamoorthy et al., 2020; Pappa, 2020; Ren et al., 2020a; Salari et al., 2020b), which have provided important initial evidence on the prevalence of mental issues at the onset of the COVID-19 crisis. One year into the COVID-19 crisis, from November 17, 2019, to November 16, 2020, we see the values of a systematic review and meta-analysis to contribute above and beyond these meta-analyses in four major directions.

First, rapid meta-analyses generally include a dozen studies (Pappa, 2020; Ren et al., 2020a; Salari et al., 2020a), most from the onset of the COVID-19 crisis; hence new systematic reviews and meta-analyses are needed to update the evidence that quickly accumulates. Our pooled prevalence rates are significantly different from, yet largely between, the findings of previous meta-analyses, suggesting our larger meta-analysis is consistent with yet revises the findings of the much smaller, previous meta-analyses. The significance of the difference between our much larger meta-analysis and the previous studies suggests a need to update meta-analyses continuously to provide more accurate estimates of the prevalence rates of mental illness during this ongoing COVID-19 epidemic.

Second, early rapid meta-analysis papers often pooled different mental disorders or distinct populations together due to the smaller numbers of studies included. However, such practices inadvertently contribute to the differences in their prevalence rates. Despite the fact that individual papers often use and report varying levels of cutoff values, most meta-analyses report the prevalence rates of mental health only by mild symptoms’ severity ^e.g., (Luo et al., 2020; Pappa et al., 2020)^. We are able to identify the major populations in the published studies (the general population, HCWs, and frontline HCWs who deal with COVID-19 patients), the major mental health outcomes (anxiety, depression, insomnia, distress, and PTSD), and the severity of outcomes (above mild, above moderate, and above severe). Moreover, we run subgroup analysis and meta-regression to reveal important differences between the mental disorders.

Third, given the large heterogeneity in terms of not only the COVID cases and deaths but also the containment strategies and hospital capacities and readiness to handle COVID-19 cases across countries(Jahanshahi et al., 2020; Zhuo et al., 2020), there are some benefits to focusing on a single country. China seems to be the first country that experienced the COVID crisis and has had a sufficient number of empirical studies to conduct such a meta-analysis.

Fourth, given our scope of the systematic review over a year of the COVID-19 crisis, our work provides a more comprehensive assessment of evidence, which is urgently needed to guide future mental health papers in the continued global pandemic. Furthermore, based on a year of mental health papers under COVID-19, we observe and provide a list of concrete issues in individual mental health papers to guide this important and proliferating stream of research.

## 2. METHODS

This systematic review and meta-analysis was conducted in accordance with the Preferred Reporting Items for Systematic Reviews and Meta-Analyses (PRISMA) statement 2019 and registered in the International Prospective Register of Systematic Reviews (PROSPERO: CRD42020220592).

### 2.1 Data Sources and Search Strategy

We conducted a comprehensive literature search in the databases of *PubMed, Embase*, and *Web of Science*. Our search query, shown in Table S1, was entered with Boolean operators to search the titles, abstracts, keywords, and subject headings (for example, Mesh terms) in each database. To account for preprints, we searched *medRxiv* (medrxiv.org). We started our search on November 10, 2020, and finalized it on November 16, 2020, one year after the first publicly known COVID-19 case (Bryner, March 14, 2020), in order to cover the first year of the COVID-19 epidemic. In addition, we checked the references of earlier rapid meta-analyses to identify other studies that may fit this review. Figure 1 details the flow chart of our search process.

**Figure 1.**
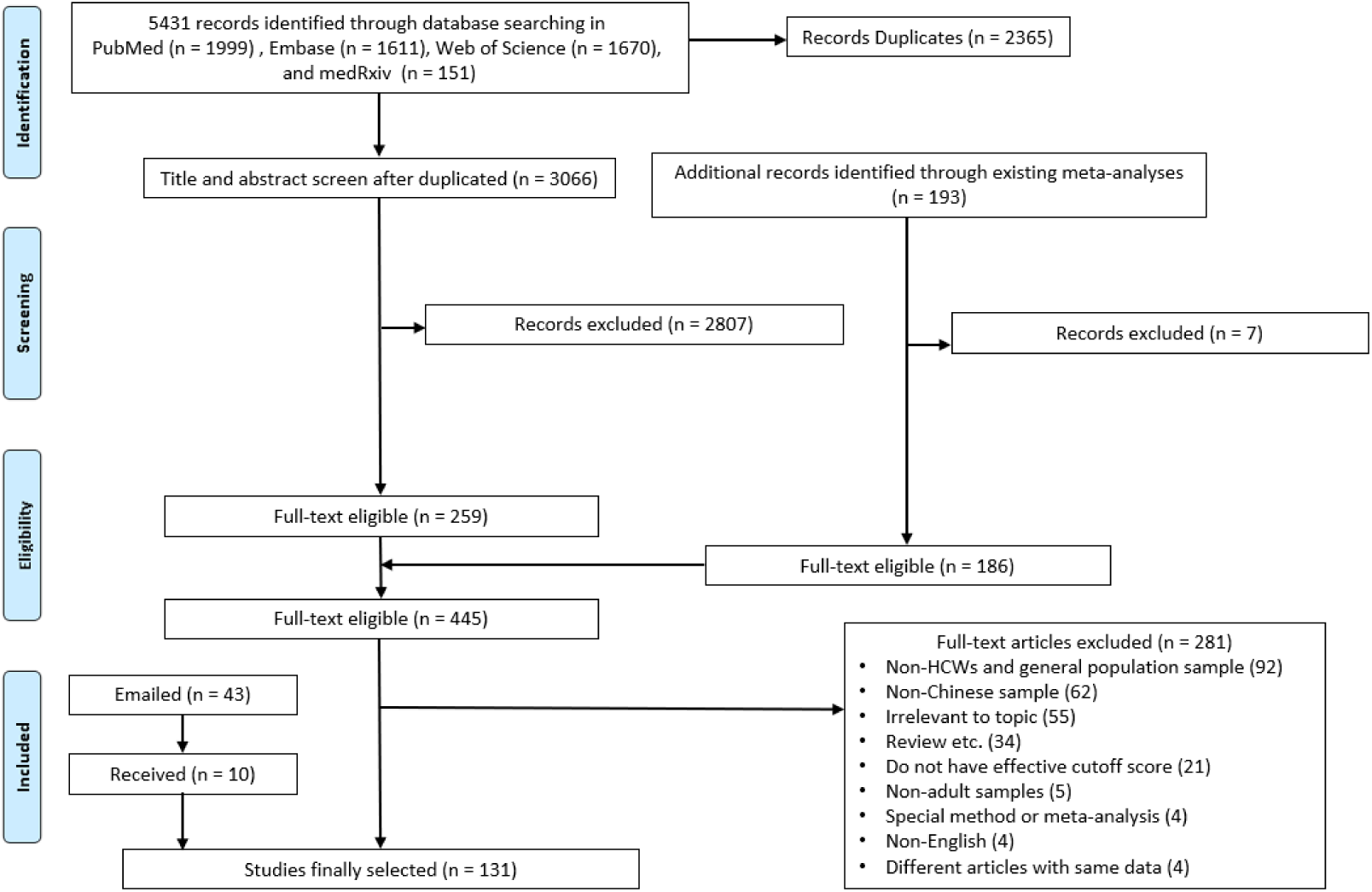
The PRISMA (Preferred Reporting Items for Systematic Reviews and Meta-Analysis) flow diagram.

### 2.2 Selection Criteria

The studies are included in our meta-analysis based on the following criteria:

a. Context: COVID-19 epidemic in China
b. Population: frontline HCWs, general HCWs, and general adult population
c. Outcome: at least one mental disorder outcomes, e.g., anxiety, depression, distress, general psychological symptoms (GPS), insomnia, and PTSD
d. Instrument: validated scales with cutoff points for the mental health outcomes
e. Language: English.

According, we excluded studies that meet the following criteria:

a. Population: children, adolescents, or specific niche adult populations such as COVID-19 patients, inpatients, or other patients, adults under quarantine, pregnant/postpartum women
b. Methodological approaches: non-primary studies such as reviews or meta-analyses, qualitative or case studies without a validated instrument, interventional studies, interviews, or news reports
c. Measurements: non-validated mental health instruments (i.e., self-made questionnaire) or instruments without a validated cutoff score to calculate a prevalence rate (i.e., STAI, SCL-90 for anxiety and depression).

We contacted the authors of papers that missed some critical information if the articles:

a. Contain primary data on mental health of relevant population using established instruments under COVID-19 period but do not report the prevalence rates. For example, a study may report the mean and SD of our outcomes but not their prevalence rates.
b. Surveyed a sample that mixed our targeted population and other populations, such as children, in a manner such that we could not extract the prevalence rate(s) for our targeted population. We included the studies that authors provided prevalence rate for our targeted population only and excluded the studies with mixed populations.
c. Miss some critical information, such as the data collection time or location.
d. Are unclear on critical information. For example, some articles are unclear whether they used the cutoff for mild or moderate symptoms to calculate the overall prevalence rates of mental issues.

### 2.3 Selection Process and Data Extraction

The articles that passed the inclusion criteria were exported into an *EndNote* library where we identified duplications and then imported to *Rayyan* for screening. Two researchers (L.T. and Y.Y.) independently screened the articles based on their titles and abstracts. If both coders excluded an article independently, it was excluded.

Six researchers (X.C, M.Z., R.C., Z.D., R.D., B.C.) were paired to assess the eligibility of each paper based on reading its full text and extracting the relevant data into a coding book based on a coding protocol. The coding book records information such as the authors and year of the paper, title, publication status, sample locations, date of data collection, sample size, response rate, population, age (mean, SD, min and max), gender proportion, instruments, cutoff scores used, the prevalence/mean/SD of the mental health outcome, and other notes or comments. Pairs of researchers first double-coded and crosschecked each paper independently. The remaining discrepancies after the crosscheck were discussed between the pair of coders. In cases where a pair of coders continued to disagree, a lead coder (X.C.) checked the paper independently and discussed it with the two original coders to determine its coding. The lead coder also integrated and reviewed all the coding information. Particularly, the lead coder checked the mental outcomes, instruments, outcome levels, and cutoff scores reported given the multitude of reporting practices in individual papers. We were able to identify papers that used unusual cutoff scores later for sensitivity analysis.

### 2.4 Assessment of Bias Risk

Following other meta-analyses (de Pablo et al., 2020; Usher et al., 2020), we used the Mixed Methods Appraisal Tool (MMAT) (Hong et al., 2018), including seven questions to conduct the quality assessment of the studies. Pairs of coders independently evaluated the risk of bias and quality of the studies and rated them based on the MMAT. Most discrepancies were resolved through a discussion between the pair of researchers, and any disagreement after discussions was resolved by a lead researcher. Papers were classed into high (6 - 7) or medium quality (lower than 6).

### 2.5 Statistical Analysis

To analyze the data in a consistent manner, we ensure the independence of mental health disorders and samples. For instance, for studies that examine a mental health outcome with more than one instrument, we report the results based on the most popular instrument. If a study reported several prevalence rates by several cutoffs, we use one of them, in the following order of preference: above the severe cutoff, above the moderate cutoff, and above the mild cutoff. Thus, only one prevalence rate for a mental health outcome in a population is entered into the meta-analysis to ensure the samples remain independent.

The overall prevalence and 95% confidence intervals of psychological outcomes were pooled using Stata 16.1. Similar to prior studies on the prevalence of mental issues, the random-effects model was used to extract the pooled estimates (DerSimonian and Laird, 1986). We reported the heterogeneity by the *I*^*2*^ statistic, which measures the percentage of variance resulting from true differences in the effect sizes rather than the sampling error (Higgins et al., 2019). We performed subgroup analyses by the key potential sources of heterogeneity of outcomes (six types of mental health disorders), severity of outcome (above mild/above moderate/above severe), and three major population groups (frontline HCWs, general HCWs, general population). Furthermore, given the high degree of heterogeneity of the true differences in the effect sizes, we ran a meta-regression to regress the prevalence upon not only these three category variables (outcome, severity, and population) but also female proportion, data collection time, data collection location (Wuhan vs. non-Wuhan), sample size, and study quality. We included data collection time to examine whether the mental issues change over time dynamically. While the COVID-19 crisis continues to evolve, there is a lack of dynamic analysis on the mental disorders of any population over time. Sensitivity analysis was conducted, and Funnel plots were used to assess publication bias. Significance level was set as two-sided and p<0.05.

## 3. RESULTS

### 3.1 Study Screening

Our systematic search (Figure 1) across all the databases yielded 5431 potentially relevant papers, out of which 2365 were duplications and removed. Of the remaining 3066 papers, we screened their titles and abstracts in the first stage and the full text of the 445 articles in the second stage. We also emailed the authors of 43 articles that missed critical information and were able to get the information to include 10 additional studies. Altogether, the process generated 131 articles for this meta-analysis (An et al., 2020; Ben-Ezra et al.; Cai et al.; Cao et al., 2020; Chen et al.; Chen et al.; Chen et al.; Chen et al.; Chen et al., 2020; Cheng et al.; Choi et al.; Dai, 2020; Dong et al., 2020; Du et al., 2020; Elhai et al., 2020; Fang et al., 2020; Feng et al., 2020; Fong et al., 2020; Fu et al., 2020; Gao, 2020; Guo, 2020; Guo et al., 2020; Han et al., 2020; Hong et al., 2020; Hou et al., 2020; Hu et al., 2020a; Hu et al., 2020b; Huang et al., 2020a; Huang et al., 2020c; Huang et al., 2020d; Jin et al., 2020; Juan et al., 2020; Lai et al., 2020; Lam et al., 2020; Lei et al., 2020; Leng et al., 2020; Li; Li et al., 2020a; Li et al., 2020b; Li et al., 2020c; Li, 2020a; Li et al., 2020d; Li, 2020b; Li et al., 2020e; Li et al., 2020f; Li et al., 2020g; Liang et al., 2020; Lin et al., 2020a; Lin et al., 2020b; Liu, 2020; Liu, in press; Liu et al., 2020a; Liu et al., 2020b; Liu et al., 2020c; Liu et al., 2020d; Liu et al., 2020e; Liu et al., 2020f; Lu et al., 2020a; Lu et al., 2020b; Lu et al., 2020c; Mi et al., 2020; Ni et al., 2020a; Ni et al., 2020b; Ning et al., 2020; Pan et al., 2020a; Pan et al., 2020b; Qi, 2020; Qian, 2020; Qian et al., 2020; Qiu et al., 2020; Que et al., 2020; Ren et al., 2020b; Shi et al., 2020; Si et al., 2020; Song et al., 2020; Song, 2020; Su et al., 2020; Sun et al., 2020a; Sun, 2020; Sun et al., 2020b; Sun et al., 2020c; Tan et al., 2020; Teng et al., 2020; Tu et al., 2020; Wang et al., 2020a; Wang et al., 2020b; Wang et al., 2020c; Wang et al., 2020d; Wang et al., 2020e; Wang et al., 2020f; Wang et al., 2020g; Wang et al., 2020h; Wang et al., 2020i; Wang et al., 2020j; Wu et al., 2020a; Wu et al., 2020b; Wu et al., 2020c; Xiao et al., 2020; Xiaoming et al., 2020; Xing et al., 2020; Xiong et al., 2020; Xu et al., 2020; Yang et al., 2020a; Yang et al., 2020b; Yin et al., 2020; Yin et al.; Ying et al., 2020; Yu et al., 2020a; Yu et al., 2020b; Zhan et al., 2020a; Zhan et al., 2020b; Zhang et al., 2020a; Zhang et al., 2020b; Zhang et al., 2020c; Zhang, 2020; Zhang et al., 2020f; Zhang et al., 2020i; Zhang et al., 2020j; Zhang et al., 2020k; Zhang and Ma, 2020; Zhang et al., 2020m; Zhao et al., 2020a; Zhao et al., 2020b; Zhao et al., 2020c; Zhou et al., 2020a; Zhou et al., 2020b; Zhou et al., 2020c; Zhu et al., 2020a; Zhu et al., 2020b; Zhu et al., 2020c; Zhu, 2020).

### 3.2 Study Characteristics

The 131 papers included contains 171 samples (Table S2) with a total of 630,244 individual participants. Table 1 summarizes their key characteristics. Among the 171 independent samples, about a quarter of them studied frontline HCWs and general HCWs (27.5% and 26.9%, respectively), and almost half studied the general population (45.6%). More than one-third of samples covered anxiety and depression. Another one-third investigated other mental issues including insomnia, PTSD, distress, and general psychological symptoms (GPS) (15.0%, 8.4%, 2.5%, and 2.0%, respectively). Respectively, 23.7%, 46.4%, and 29.9% of samples reported prevalence rates at the mild above, moderate above, and severe above level by the severity of the symptoms.

**Table 1.** **Characteristics of the studies on mental health in China in a year of COVID-19 epidemic**

| Characteristics | Total number of studies/samples | Percent | Level of analysis |
| --- | --- | --- | --- |
| *Overall* | 131 | 100 | Article |
| *Population* |  |  | Sample |
| Frontline HCWs | 47 | 27.5 |  |
| General HCWs | 46 | 26.9 |  |
| General population | 78 | 45.6 |  |
| *Outcome* |  |  | Prevalence |
| Anxiety | 127 | 35.5 |  |
| Depression | 128 | 35.8 |  |
| Distress | 9 | 2.5 |  |
| General psychological symptoms | 7 | 2.0 |  |
| Insomnia | 57 | 15.9 |  |
| PTSD | 30 | 8.4 |  |
| *Severity* |  |  | Prevalence |
| Above mild | 85 | 23.7 |  |
| Above moderate | 166 | 46.4 |  |
| Above severe | 107 | 29.9 |  |
| *Sampling location* |  |  | Article |
| Wuhan | 38 | 22.2 |  |
| Non-Wuhan | 123 | 77.8 |  |
| *Sampling date* |  |  | Article |
| January 2020 | 9 | 6.9 |  |
| February 2020 | 85 | 64.9 |  |
| March 2020 | 23 | 17.6 |  |
| April 2020 | 8 | 6.1 |  |
| May 2020 | 2 | 1.5 |  |
| June 2020 | 2 | 1.5 |  |
| July 2020 | 2 | 1.5 |  |
| *Design* |  |  | Article |
| Cross-sectional | 126 | 96.2 |  |
| Cohort | 5 | 3.8 |  |
| *Publication status* |  |  | Article |
| Preprint | 10 | 7.6 |  |
| Accepted | 1 | 0.8 |  |
| Published | 120 | 91.6 |  |
| *Quality* |  |  | Article |
| Good | 100 | 73.3 |  |
| Medium | 31 | 23.7 |  |
|  | Median | Range |  |
| *Number of participants* | 709 | 30 - 123,768 | Article |
| *Female portion* | 69% | 12% - 100% | Article |
| *Response rate* | 85% | 14% - 100% | Article |

Almost all the studies, 126 out of 131, employed cross-sectional surveys; specifically, 9 (6.9%) conducted the survey in January 2020, 85 (64.9%) in February, 23 (17.6%) in March, and 14 (10.6%) in April or later. Almost one-quarter of them (22.2%) contained a sample targeting populations in Wuhan. Most studies were published in journals, and 10 (7.6%) studies remained as preprints. The assessment based on the Mixed Methods Appraisal Tool (MMAT) indicated 100 (73.3%) studies were of good quality (score no less than 6 out of 7) and 31 studies were of medium quality (score less than 6 but greater than 4). The median number of individuals per sample was 709 (range: 30 to 123,768) with a median female proportion of 69% (range: 12% to 100%) and a median response rate of 5% (range: 14% to 100%).

The 131 papers employed a wide arrange of instruments to assess mental health (Table S3). For both anxiety and depression, PHQ (60.6%, 63.3%) and SAS (23.6%, 13.3%) are the first and second most popular measures; distress is measured the most by K6 (44.4%), followed by IES-R (22.2%); insomnia is measured by ISI (63.2%) and PSQI (29.8%); PTSD by IES-R (40.0%), PCL-C (26.7%), and PCL-5 (26.7%); and general psychological symptoms by SRQ-20 (100.0%). Please see the details in Table S3.

### 3.3 Major Issues from Findings of the Key Study Characteristics

Our systematic review reveals several widespread issues in mental health research during COVID-19: a wide array of instruments, inconsistent reporting of prevalence rates, inconsistent use and reporting of cutoff points, different cutoff values to determine the overall prevalence as well as the severity, and other issues on reporting standards and terminologies.

#### A myriad of instruments

The individual papers on mental health research during COVID-19 employed a wide variety of instruments with varying degrees of popularity and validity (summarized in Table S3). The wide array of instruments, especially the use of less frequently used instruments (i.e., AIS, BAI/BDI), certainly has some benefit but makes it hard to make comparisons or accumulate evidence.

#### Admixed outcome severity level

The individual papers reported the prevalence rates at a range of severity of the symptoms. First, the articles differ in their terminology when reporting the overall prevalence rates. Some papers used the overall prevalence rate to indicate the percentage with moderate symptoms or above, other papers used it to indicate the percentage with mild symptoms or above ^e.g., (Du et al., 2020) (Zhang et al., 2020b)^. Even worse, a large number of papers did not specify the definition of the overall prevalence rate, rendering it impossible to know whether it refers to above mild or moderate levels. Second, some papers use other terminologies, such as “extremely severe” (Ozamiz-Etxebarria et al., 2020), “very severe” (Moghanibashi-Mansourieh, 2020), or “very high” (Temsah, 2020), “moderate-severe” ^e.g.,^ (Guiroy et al., 2020), “moderate to severe” (Moccia et al., 2020; Wang et al., 2020a), “moderately severe” ^e.g.,^ (Xiaoming et al., 2020), and “poor” (Wang et al., 2020f), which often is not clear in terms of the cutoff points used to categorize those symptoms. We opted to recode all the papers that indicate their cutoff scores manually(Antony et al., 1998; Beusenberg et al., 1994; Blevins et al., 2015; Buysse et al., 1989; Cheng et al., 2002; Cheung et al., 2007; Creamer et al., 2003; Dai, 2020; Dobie et al., 2002; Guo et al., 2017; Health; Kessler et al., 2010; Kroenke et al., 2001; Kroenke et al., 2009; Lee et al., 2018; Matza et al., 2010; Morin et al., 2011; Prins et al., 2016; Qiu et al., 2020; Schlenger et al., 2002; Soldatos et al., 2000; Spitzer et al., 2006; Thoresen et al., 2010; Tsai et al., 2005; Wang et al., 2017; Wang et al., 2011; Wilberforce et al., 2010; Wu et al., 2003; Zigmond and Snaith, 1983; Zimmerman et al., 2013; Zung, 1965, 1971; 王春芳 et al., 1986); however, these terminologies may contribute to the heterogeneity and confusion in accumulating evidence.

#### Clarity on the cutoff points used to determine severity

Some papers employed nonstandard or unusual cutoff scores ^e.g., (Elhai et al., 2020)^, at times without referencing validation studies that supported the use of those special cutoff scores ^e.g., (Cai et al.; Song et al., 2020)^. Some papers did not report the cutoff score used or provide any references ^e.g., (Cao et al., 2020; Sun et al., 2020b)^, making the assessment difficult. Such issues seem to occur particularly in studies that used PSQI, IES-R and DASS-21, and CES-D.

### 3.4. Pooled Prevalence Rates of Mental Health Disorders

The prevalence rates of the 171 samples were pooled by the subgroups (Table 2). First, the overall prevalence rates of mental health disorders that surpassed the cutoff values of mild, moderate, and severe symptoms were 27%, 18%, and 3%, respectively. The overall prevalence of mental health disorder frontline HCWs, general HCWs, and the general population in China are 17%, 14%, and 12%, respectively. The overall prevalence of anxiety, depression, distress, GPS, insomnia, and PTSD are 11%, 13%, 20%, 13%, 19%, and 20%. Figure 2 graphically depicts such findings of the pooled analysis by subgroups using forest plots.

**Table 2.**
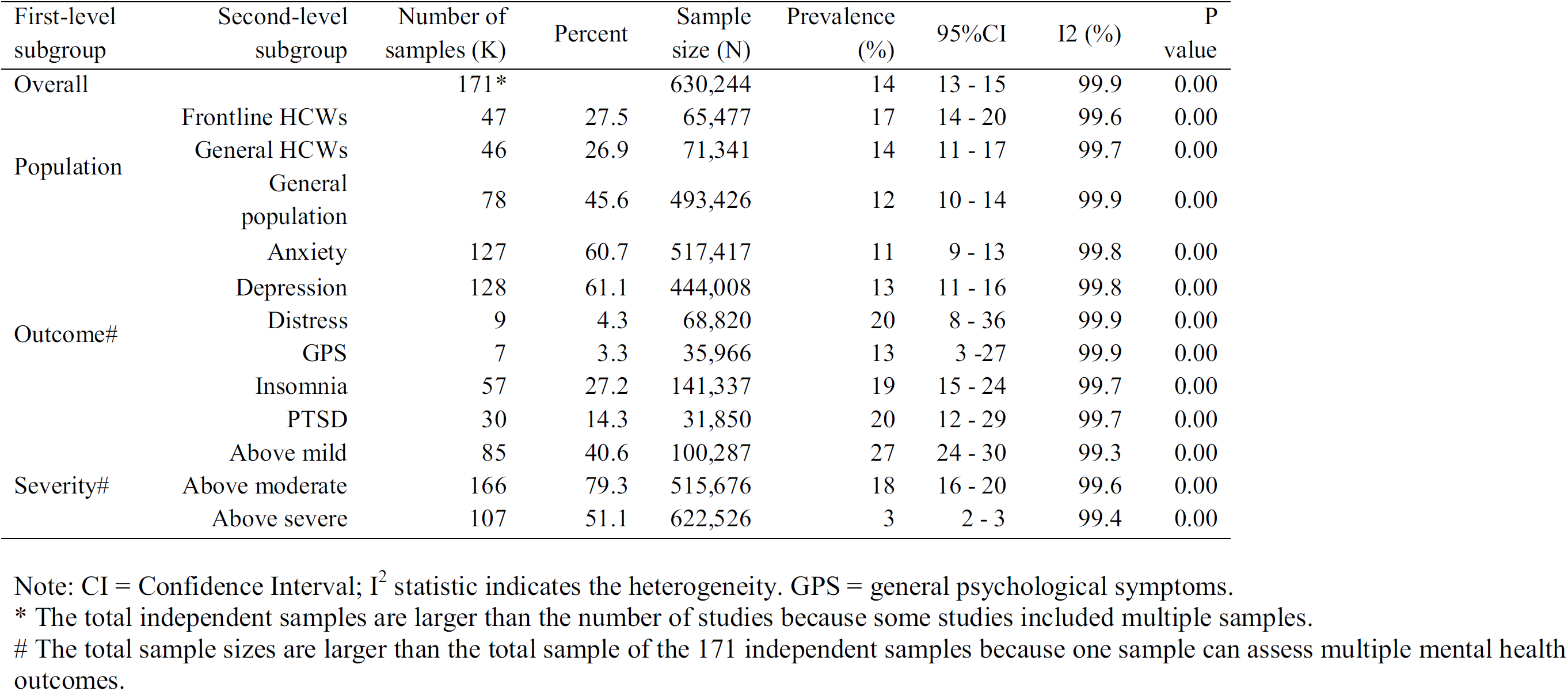
The pooled prevalence rates of mental health disorders by subgroups of population, outcome, and severity

**Figure 2a.**
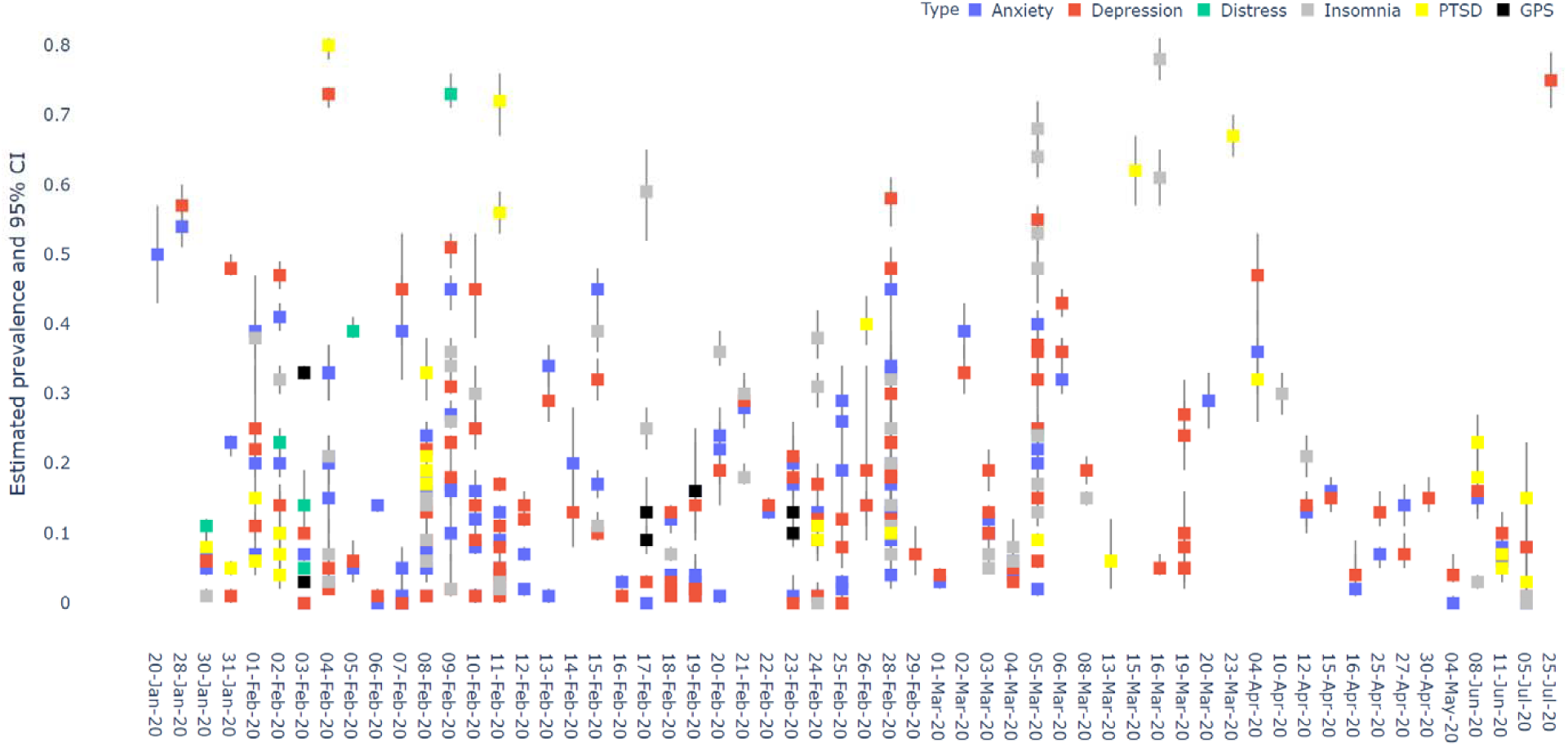
A forest plot of the pooled prevalence by outcomes.

**Figure 2b.**
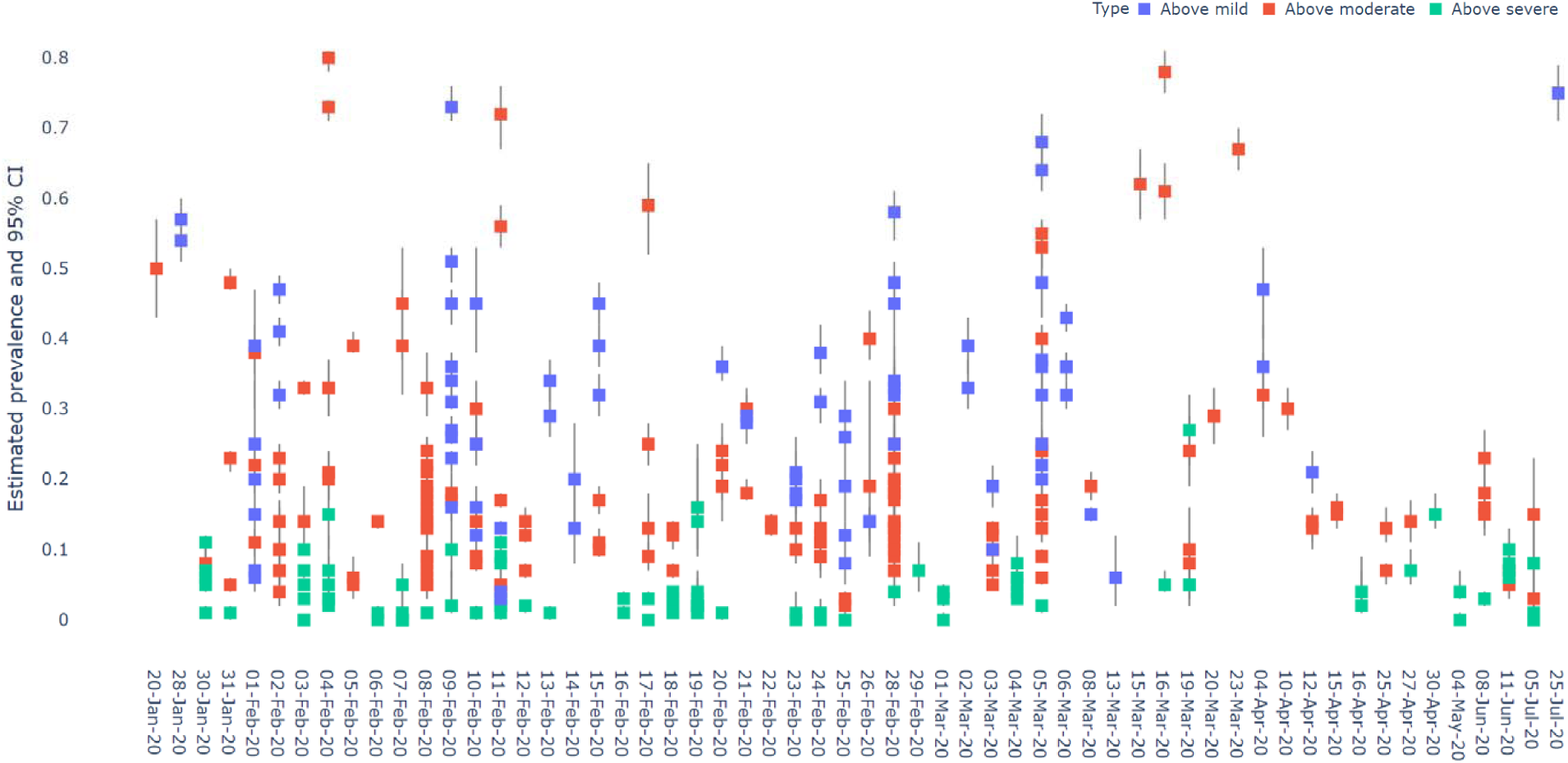
A forest plot of the pooled prevalence by outcome levels.

**Figure 2c.**
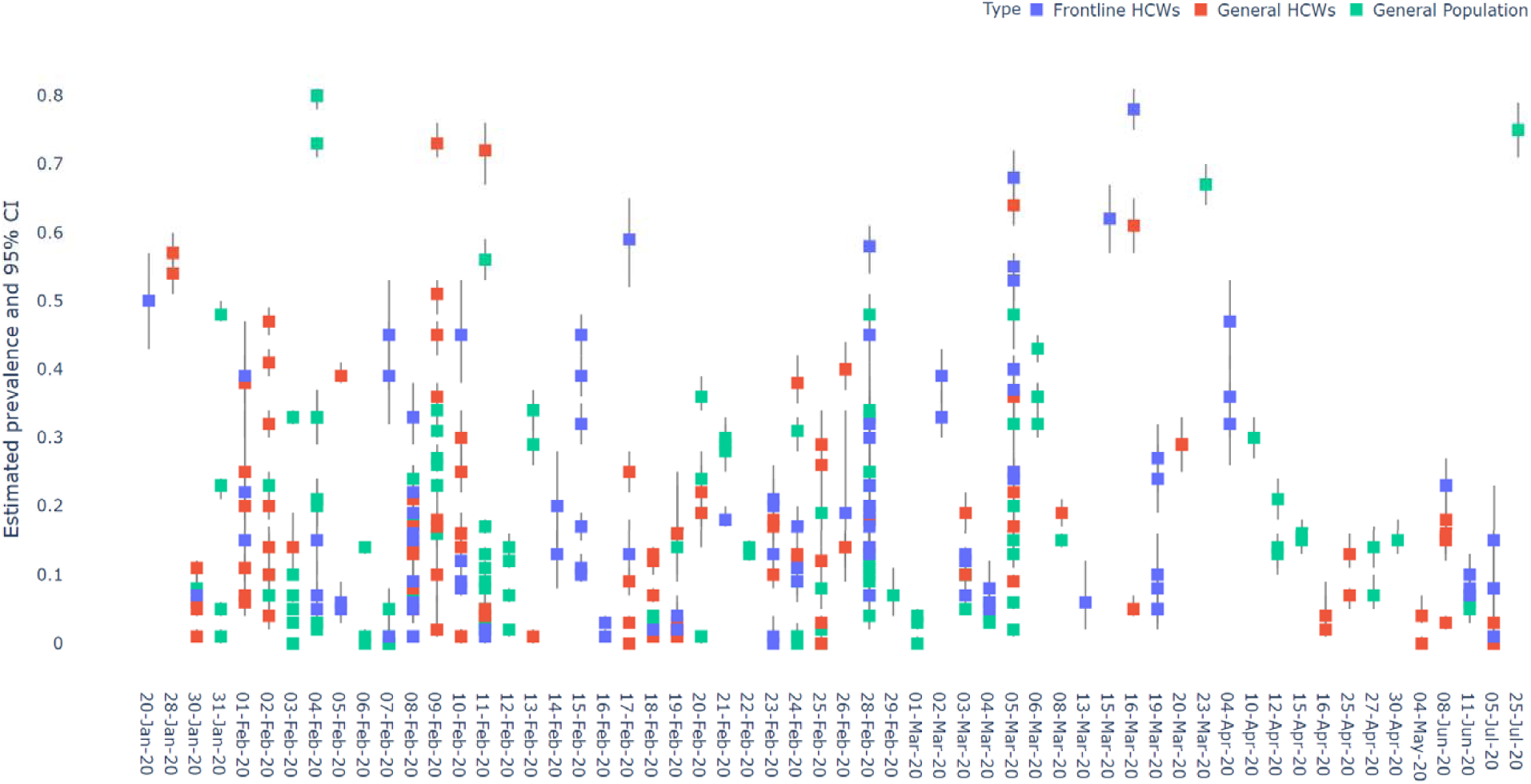
A forest plot of the pooled prevalence by population.

### 3.5 Meta-regression on the Prevalence of Mental Health Disorders

To better explain the heterogeneity of the prevalence of mental health disorders, Table 3 reports the results of a meta-regression analysis. The meta-analytical model explained over 40% of the variance of mental health disorders among these studies (*R-squared* = 41.0%, *I*^*2*^ = 100%, *tau*^*2*^ = 0.11). The prevalence of severe mental health disorders is significantly lower than that of moderate mental illness (*p*<0.01), which is in turn significantly lower than those of mild mental illness (*p*<0.01). The prevalence of mental health disorders of frontline HCWs is significantly higher than that of general HCWs (*p*<0.004). General HCWs and the general population do not differ in their mental health prevalence rates. The prevalence rates of depression (*p*=0.04) and insomnia (*p*=0.04) are significantly higher than that of anxiety, and the rates of general psychological symptoms (*p*=0.20) and PTSD (*p*=0.20) do not differ significantly from that of anxiety. Interestingly, the prevalence of mental health disorders of participants in Wuhan, the epicenter of the COVID-19 crisis, was significantly lower than that in non-Wuhan samples in China (*p*=0.04). The prevalence rates of mental health disorders were higher in studies of papers with a higher quality rating (*p*=0.03). The female proportion (*p*=0.54), date of data collection (*p*=0.64), sample size of studies (*p*=0.16), or publication status (*p*=0.80) did not predict the prevalence rates significantly.

**Table 3.** **The results of meta-regression of mental health disorders during COVID-19**

| Variables | Coefficient (CI, 95%) | Std. Err. | P-value |
| --- | --- | --- | --- |
| *Outcome* |  |  |  |
| Anxiety (reference) |  |  |  |
| Depression | 0.03 (-0.06 to 0.11) | 0.04 | 0.55 |
| Distress | 0.24* (0.01 to 0.47) | 0.12 | 0.04 |
| General psychological symptoms | -0.01 (-0.27 to 0.26) | 0.14 | 0.97 |
| Insomnia | 0.12* (0.01 to 0.23) | 0.06 | 0.04 |
| PTSD | 0.10 (-0.05 to 0.24) | 0.07 | 0.20 |
| *Severity* |  |  |  |
| Above mild | 0.20*** (0.11 to 0.30) | 0.05 | 0.00 |
| Above moderate (reference) |  |  |  |
| Above severe | -0.49*** (-0.58 to 0.40) | 0.05 | 0.00 |
| *Population* |  |  |  |
| Frontline HCWs | 0.14*** (0.05 to 0.23) | 0.05 | 0.00 |
| General HCWs (reference) |  |  |  |
| General population | 0.03 (-0.07 to 0.13) | 0.05 | 0.51 |
| *Publication Status* |  |  |  |
| Preprint (reference) |  |  |  |
| Accepted | -0.34 (-0.84 to 0.17) | 0.26 | 0.19 |
| Published | -0.02 (-0.17 to 0.13) | 0.08 | 0.80 |
| *Female proportion* | 0.09 (-0.19 to 0.37) | 0.14 | 0.54 |
| *Date of data collection* | 0.00 (0.00 to 0.00) | 0.00 | 0.64 |
| *Wuhan vs. Non-Wuhan sample* | -0.10* (-0.20 to 0.00) | 0.05 | 0.04 |
| *Sample size* | 0.00 (0.00 to 0.00) | 0.00 | 0.16 |
| *Quality* | 0.08* (0.01 to 0.15) | 0.04 | 0.03 |
| Constant | 5.99 | 12.36 | 0.63 |
| R<sup>2</sup> | 0.41 |  |  |
| Wald X<sup>2</sup> (16) | 252.05*** |  | 0.00 |

The meta-analytical results enable the prediction of prevalence rates while taking account of the influence of multiple factors and hence offer a superior model over the earlier pooled analyses. In other words, the meta-regression model considers multiple predictors of mental health disorders in a single model at the same time instead of the approach of considering one predictor at a time by pooled prevalence, the typical method to estimate the prevalence of mental health disorder in prior meta-analytical papers in COVID-19 literature. Hence, based on the results of the meta-regression, we report the predicted prevalence rates of varying severity levels of the symptoms of different mental health disorders of frontline HCWs, general HCWs, and the general population. Table 4 and Figure 3 show the predicted prevalence rates of mental health disorders by populations, outcomes, and severity by the meta-analytical regression model. As illustrated in Figure 3, the prevalence rates vary greatly by the mental health outcomes and severity. The prevalence rates are lower when using a higher level of severity, which drives the heterogeneity of prevalence rate to a large degree. Among the different types of mental health outcomes, distress seems to be the most prevalent among all three populations.

**Table 4.**
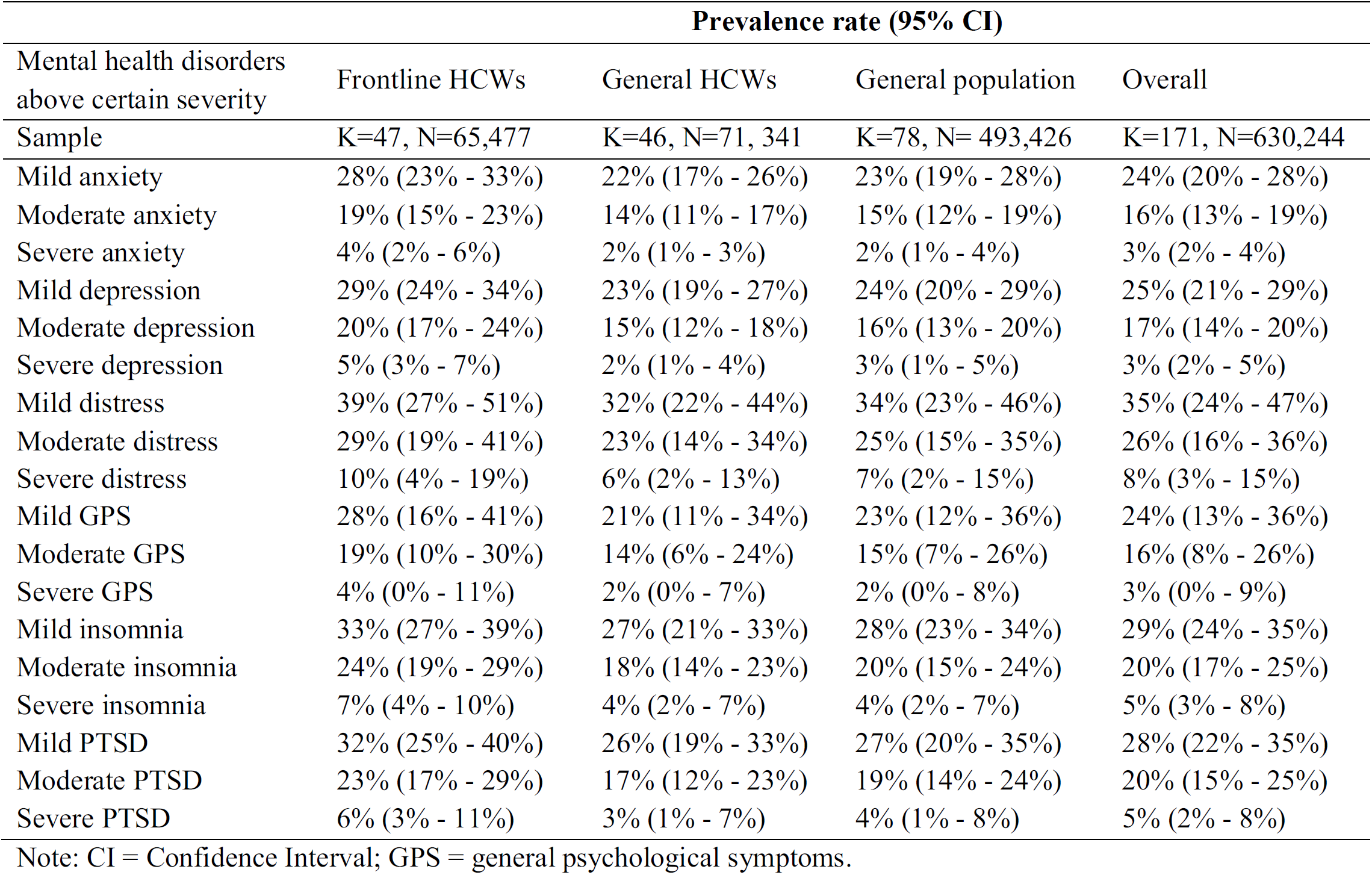
**The predicted prevalence rates of mental health disorders by populations, outcomes, and severity by the meta-analytical regression model**

| Mental health disorders<br>above certain severity | Prevalence rate (95% CI) |  |  |  |
| --- | --- | --- | --- | --- |
|  | Frontline HCWs | General HCWs | General population | Overall |
| Sample | K=47, N=65,477 | K=46, N=71, 341 | K=78, N= 493,426 | K=171, N=630,244 |
| Mild anxiety | 28% (23% - 33%) | 22% (17% - 26%) | 23% (19% - 28%) | 24% (20% - 28%) |
| Moderate anxiety | 19% (15% - 23%) | 14% (11% - 17%) | 15% (12% - 19%) | 16% (13% - 19%) |
| Severe anxiety | 4% (2% - 6%) | 2% (1% - 3%) | 2% (1% - 4%) | 3% (2% - 4%) |
| Mild depression | 29% (24% - 34%) | 23% (19% - 27%) | 24% (20% - 29%) | 25% (21% - 29%) |
| Moderate depression | 20% (17% - 24%) | 15% (12% - 18%) | 16% (13% - 20%) | 17% (14% - 20%) |
| Severe depression | 5% (3% - 7%) | 2% (1% - 4%) | 3% (1% - 5%) | 3% (2% - 5%) |
| Mild distress | 39% (27% - 51%) | 32% (22% - 44%) | 34% (23% - 46%) | 35% (24% - 47%) |
| Moderate distress | 29% (19% - 41%) | 23% (14% - 34%) | 25% (15% - 35%) | 26% (16% - 36%) |
| Severe distress | 10% (4% - 19%) | 6% (2% - 13%) | 7% (2% - 15%) | 8% (3% - 15%) |
| Mild GPS | 28% (16% - 41%) | 21% (11% - 34%) | 23% (12% - 36%) | 24% (13% - 36%) |
| Moderate GPS | 19% (10% - 30%) | 14% (6% - 24%) | 15% (7% - 26%) | 16% (8% - 26%) |
| Severe GPS | 4% (0% - 11%) | 2% (0% - 7%) | 2% (0% - 8%) | 3% (0% - 9%) |
| Mild insomnia | 33% (27% - 39%) | 27% (21% - 33%) | 28% (23% - 34%) | 29% (24% - 35%) |
| Moderate insomnia | 24% (19% - 29%) | 18% (14% - 23%) | 20% (15% - 24%) | 20% (17% - 25%) |
| Severe insomnia | 7% (4% - 10%) | 4% (2% - 7%) | 4% (2% - 7%) | 5% (3% - 8%) |
| Mild PTSD | 32% (25% - 40%) | 26% (19% - 33%) | 27% (20% - 35%) | 28% (22% - 35%) |
| Moderate PTSD | 23% (17% - 29%) | 17% (12% - 23%) | 19% (14% - 24%) | 20% (15% - 25%) |
| Severe PTSD | 6% (3% - 11%) | 3% (1% - 7%) | 4% (1% - 8%) | 5% (2% - 8%) |

Note: CI = Confidence Interval; GPS = general psychological symptoms.

**Figure 3.**
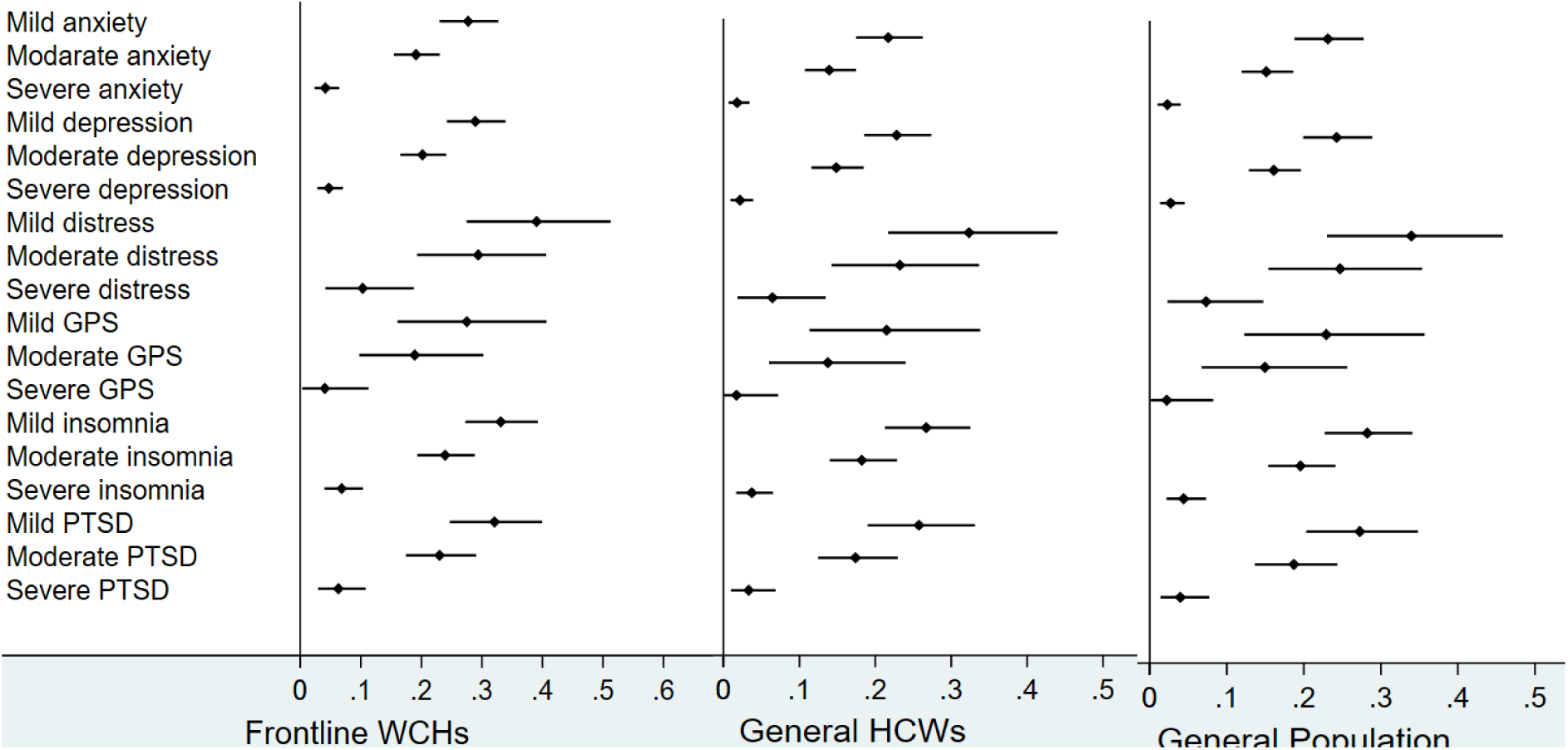
The predicted prevalence rates of mental health disorders by populations, outcomes, and severity by the meta-analytical regression model.

### 3.6 Sensitivity Analysis

Our meta-analytical model was able to take account of the impact of several factors, such as publication status (insignificant), sample size (insignificant), and article quality score (significant). Furthermore, we conducted our analysis with the exclusion of each study one-by-one from the meta-analytic model and found it did not significantly alter the findings. The visual inspection of the sensitivity plot however revealed that there is significant asymmetry. Figure 4 reports the DOI plot in combination with the Luis-Kanamori (LFK) index, which has higher sensitivity and power than a funnel plot (Mboua et al., 2020; Teshome et al., 2020). An LFK index scores of ±1, between ±1 and ±2, or ±2 indicating ‘no asymmetry’, ‘minor asymmetry’, and ‘major asymmetry’ respectively, and hence the LFK index of 2.1 represents major asymmetry. Therefore, the presence of publication bias is likely.

**Figure 4.**
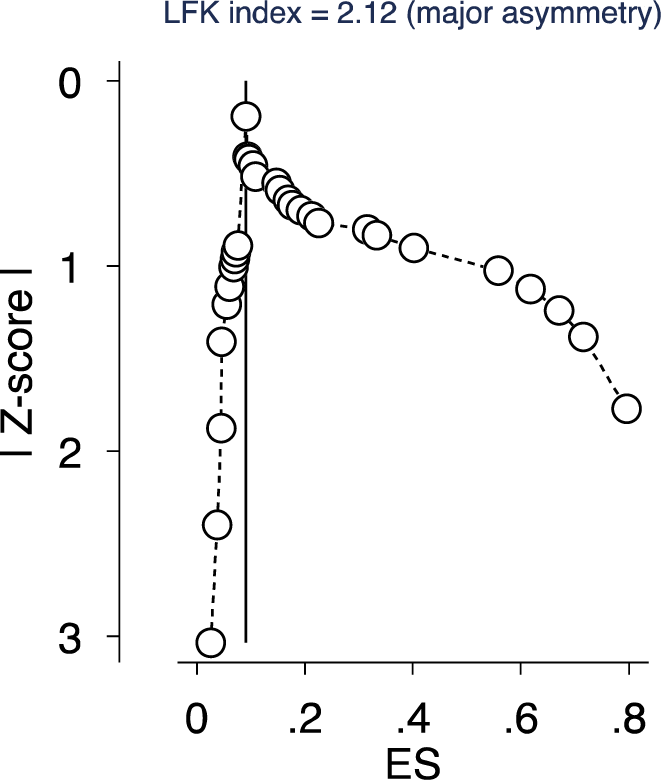
The DOI plot and the Luis Furuya–Kanamori (LFK) index over ±2 indicates ‘major asymmetry’ in publication bias among the studies published to date.

## 4. DISCUSSION

### 4.1 Comparison with Prior Meta-analyses

The meta-analysis of mental health one year into the COVID-19 epidemic in China revealed several findings that are worth comparing with prior meta-analyses on the same topic. See Table 5 for a summary of the comparison.

**Table 5.**
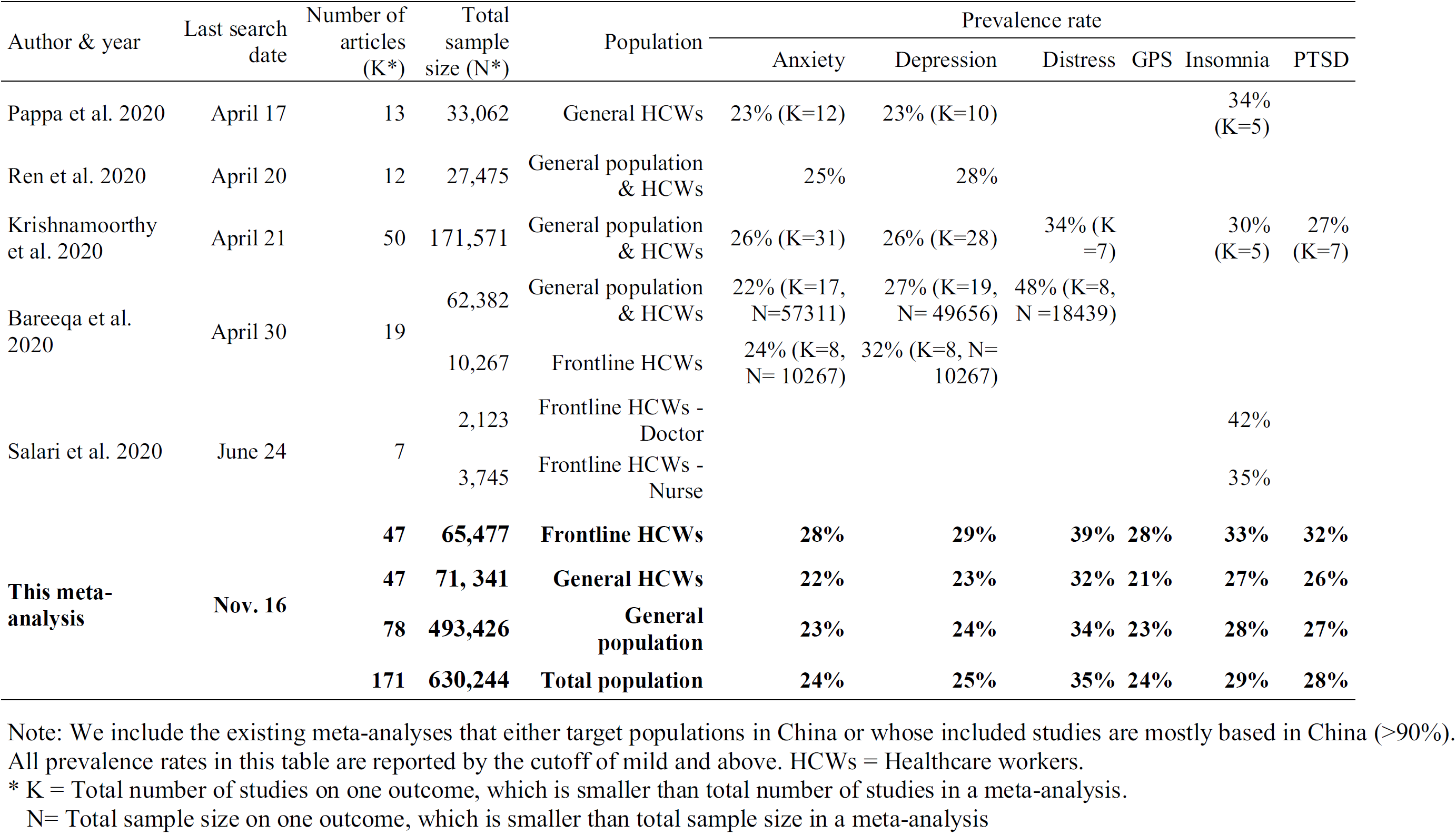
**Comparisons with prior meta-analyses of similar topic**

Unlike prior meta-analyses, most of which searched the literature before May 2020, our meta-analysis covered a whole year of COVID-19 to yield stronger evidence. Our meta-analysis from a systematic review comprises 171 independent samples with 630,244 participants from 131 studies, much larger than the prior meta-analyses on China’s population that included 7–50 studies with 2123 to 62,382 participants (Bareeqa et al., 2020; Krishnamoorthy et al., 2020; Pappa, 2020; Ren et al., 2020a; Salari et al., 2020b). The comparison reveals that our pooled prevalence rates largely fall between the findings of previous meta-analyses, suggesting our larger data is consistent yet fine-tunes them. For example, we reported a higher prevalence of anxiety for the general population and HCWs (24%) than Bareeqa et al. (2020) (22%) and Pappa et al. (2020) (23%), but lower than Krishnamoorthy et al. 2020 (26%) and Ren et al. 2020 (25%). Similarly, we reported a higher prevalence of anxiety for frontline HCWs (28%) than Bareeqa et al. (2020) (24%) and a lower prevalence of depression (25%) for the general population and HCWs than the prevalent rates of 26% - 28% in Ren et al. 2020, Krishnamoorthy et al. 2020, and Bareeqa et al. 2020. All these differences between our prevalence rates and the prior reports (Bareeqa et al., 2020; Krishnamoorthy et al., 2020; Pappa, 2020; Ren et al., 2020a) are statistically significant due to the large sample size involved, and hence we significantly update the cumulative evidence on mental health prevalence rates in COVID-19. Our findings also suggest a need to update meta-analyses continuously to provide more accurate estimates of the prevalence of mental illness while COVID-19 is ongoing.

Our systematic review over a year of the COVID-19 crisis allows us to identify all the major mental health outcomes studied (anxiety, depression, insomnia, GPS, distress, and PTSD). In particular, GPS has never been included in any prior meta-analysis. Moreover, prior meta-analyses examined the prevalence rates of mental health disorders based on one level of the severity of symptoms (i.e., above mild), and we included articles that reported the prevalence at varying levels of severity of symptoms.

### 4.2 Meta-regression Findings

Thanks to the large number of samples in China over a year of the COVID-19 crisis, we were able to conduct meta-regression to account for the influence of multiple predictors at the same time to enable better prediction on the prevalence of each mental health disorder. The accumulative evidence shows that several predictors are significantly associated with prevalence rates of mental issues in China during COVID-19, including the severity and type of mental issues, population, sampling location, and study quality.

The severity of mental symptoms, which has been unaccounted for in prior meta-analyses, was found to contribute greatly to the heterogeneity of prevalence rates, hence individual mental health papers need to pay special attention to the severity with clarity. Otherwise, researchers and practitioners might mix the severity of severe, moderate, and mild mental illness. Since prior meta-analyses largely examined the prevalence rates of mild mental health disorders, yet psychiatrists care not only the mild symptoms, and the significant differences revealed by this study call for more meta-analyses on varying levels of severity to provide evidence for practitioners relevant to their concerns.

Among the six types of mental health issues examined, distress and insomnia had the highest prevalence rates among all three populations. Our findings suggest that practitioners need to be aware and pay more attention to distress and insomnia under the COVID-19 pandemic. Moreover, given that more than two-thirds of existing empirical studies focused on anxiety and depression, we call out for future research to focus on mental distress and insomnia.

Frontline HCWs suffered more than general HCWs and the general population across all six types of mental issues. It is also worth noting the general HCWs did not significantly differ from general populations across any mental issues. Hence, our evidence suggests that policymakers and healthcare organizations need to further prioritize frontline HCWs the most in this ongoing pandemic.

Past mental health research has reported inconsistent results on the relationship between individuals’ mental issues and their locations. Some studies reported that mental issues increase along with the distance to the epicenter in the COVID-19 pandemic, known as “typhoon eye effect” (Tang et al., 2020; Zhang et al., 2020d; Zhang et al.). However, other findings have demonstrated an opposite effect, where mental issues decrease as the distance to the epicenter increases, known as the “ripple effect” (Huang et al., 2020b; Zhang et al., 2020l). Our accumulative evidence shows that people in the epicenter of China in Wuhan suffered less mental issues than those outside of Wuhan, lending support to the typhoon eye effect. This finding suggests future research to differentiate, report, and possibly model sampling locations based on the epicenter of a pandemic to enable better geographical identification of mental issues (Yáñez, 2020; Zhang et al., 2020e; Zhang et al., 2020g).

Our findings that the samples in papers with higher quality tend to find higher prevalent rates of mental issues suggest study quality may matter. Particularly, future meta-analysis may pay attention to the representativeness of sampling, the response rate, etc., to better account for the heterogeneity in the pooled prevalence rates.

As the COVID-19 epidemic evolves, we expected the mental issues may change over time. However, the evidence of meta-regression using time as a predictor failed to reveal significant effect, even though the COVID crisis has evolved to varying degrees for more than a year in countries such as China. A potential reason might be the development of COVID-19 in various parts of China happened at varying paces, and more refined studies are needed to uncover the change of prevalence rates effect over time.

### 4.3 A Mental Health Research Agenda during Covid-19

Our systemic review and meta-analysis allowed us to observe several widespread problems in the individual papers that impede the accumulation of evidence. We offer a few concrete suggestions on research focus and reporting for future mental health studies for authors, editors, and reviewers in a table for easy reference (Table 6) to improve the quality of such studies and to facilitate evidence accumulation in future meta-analyses.

**Table 6.** **A list of recommendations for mental health research papers**

|  |  |
| --- | --- |
|  | Guides for future research and reporting |
| Outcome and instrument | 1) Study health outcomes that have higher prevalence rates, e.g., distress and insomnia<br>2) Use validated instruments |
| Severity of the symptoms | 3) Use and report more levels of severity of symptoms and the cutoff points used<br>4) Specify the meaning of overall prevalence, whether above mild or above moderate<br>5) Specify the cutoff values used with the reasons/references |
| Characteristics of the samples | 6) Report sampling locations more precisely – not just the country, but the region or the distance from epicenter if possible<br>7) Report the sampling dates<br>8) Report the age/gender of the participants |
| Population | 9) Separate and focus on frontline line HCWs vs. HCWs |
| Study design | 10) More future research using cohort designs |

### 4.4 Study Limitations

This meta-analysis has a few limitations. First, the validity of our findings rests upon the quality and reporting of the original studies. As discussed before, individual mental health papers varied in their usage of instruments, cutoff scores, the use of cutoff scores to define mental issues, and the reporting standards. For example, the overall prevalence refers to “above the cutoff of mild” in some papers yet “above the cutoff of moderate” in other papers. Worse, many papers report the overall prevalence without specifying which/how cutoff scores are used. While we paid extra attention to the severity, the cutoff points, and the ways in which individual articles used this information, the multitude of varying practices contributes to additional noise and variance in the analysis. Second, since we included studies in English, which may result in some biases. Third, 96.2% of studies included in this meta-analysis were cross-sectional surveys, and we call for more cohort studies to examine the effect of time. Finally, we only focus on studies that collected data in China, and we call for future meta-analyses in other countries or regions as the COVID-19 crisis continues in most parts of the world.

### 4.5 Conclusion

Since the COVID-19 epidemic started in November 2019, hundreds of studies have documented the mental health of major populations by the key mental outcomes and varying levels of severity. This systematic review and meta-analysis synthesized the evidence on the prevalence rates of mental health disorders in China over one year of the COVID-19 epidemic. The meta-regression results provide evidence that future research should pay attention to mental distress and insomnia, especially given the popularity of anxiety and depression in the literature to date. Moreover, we revealed a number of issues in the individual papers published on mental health during the COVID-19 crisis, and the high heterogeneity among studies calls for more standard reporting of future research not only during COVID-19 but also generally to better facilitate the synthesis of evidence to enable evidence-based research and practice.

## Data Availability

All data generated or analyzed during this study are included in this published article.

## Credit author statement

XC: *Investigation, Data curation, Visualization, Writing – original draft, Writing – review & editing, Project administration*. JC: *Conceptualization, Methodology, Validation, Formal analysis, Investigation, Resources, Data curation, Visualization, Writing – original draft, Writing – review & editing, Supervision*. MZ, RC, RD, ZD, YY, BC, LT: *Investigation (Data)*. RZ, WC, PL: *Investigation*. SZ: *Conceptualization, Methodology, Validation, Formal analysis, Investigation, Data curation, Writing – original draft, Writing – review & editing, Supervision*. XC, JC, and SZ co-lead this project. All authors were involved in approving the manuscript. The corresponding author attests that all listed authors meet authorship criteria and that no others meeting the criteria have been omitted.

## Transparency declaration

*The lead author* affirms that this manuscript is an honest, accurate, and transparent account of the study being reported; that no important aspects of the study have been omitted; and that any discrepancies from the study as planned (and, if relevant, registered) have been explained*.

## Ethical appr oval

*Not applicable*

## Patient and public involvement

*No patient or public was involved in a systematic review and meta-analysis*

## Acknowledgements

*We thank Haixing Zheng, Shaokun Xu, Fei Liang & Zhehong Xu for their help*.

## Data sharing statement

*All data generated or analyzed during this study are included in this published article*.

