## Supplementary materials table S1 - S3 for "One Year of Evidence on Mental Health Disorders in China during the COVID-19 Crisis - A Systematic Review and Meta-Analysis"

**Table S1: The search string used in this systematic review and meta-analysis**
(Nov 17, 2019 - Nov 16, 2020)

| Search | Search topic | Search keywords (titles, abstracts, and subject headings) with Boolean operators |
|---|---|---|
| 1 | Exposure/ Context | "coronavirus" OR "SARS-COV-2" OR "COVID-19" OR “2019nCoV” OR “2019-nCoV” |
| 2 | Outcome of interest | "depressi*" OR "anxi*" OR "insomnia" OR "sleep disorder" OR "sleep issue" OR "sleep problem" OR "PTSD" OR "distress disorder" OR "distress symptom" OR "distress issue" OR "distress problem" OR "mental disorder" OR "mental issue" OR "mental problem" OR "psychiatric disorder" OR "psychiatric issue" OR "psychiatric problem" |
| 3 | Language | English |
| Overall search | | 1 AND 2 AND 3 |

### Table S2. The characteristics of the individual studies included

| Authors, year | Population | Sample size | Wuhan vs. Not | Female proportion | Outcome (%) | | | | | | Instrument | Outcome Severity |
|---|---|---|---|---|---|---|---|---|---|---|---|---|
| | | | | | ANX | DEP | DIS | GPS | INS | PTSD | | |
| An et al., 2020 | GHCW | 1103 | No | 90.7 | NA | 5.3 | NA | NA | NA | NA | PHQ-9 | Severe |
| Ben-Ezra et al., 2020 | GP | 1134 | No | 53.5 | NA | NA | NA | 19.1 | NA | NA | K-6 | Moderate |
| Cai et al., 2020 | FHCW | 348,<br>343 | Yes | 96.5<br>97.4 | 16.4<br>4.6 | 21.8<br>9.2 | 14.7<br>6.1 | NA | 33.3<br>19.1 | NA | GAD-7, PHQ-9, ISI, IES-R | Moderate |
| | GHCW | 361<br>278 | | | 4.7<br>7.6 | 9.0<br>12.8 | 9.0<br>13.7 | NA | 16.9<br>21.3 | NA | | |
| Cao et al., 2020 | FHCW | 37 | Yes | 78.4 | NA | 18.9 | NA | NA | NA | NA | PHQ-9 | Moderate |
| Chen et al., 2020a | GHCW | 105 | No | 90.5 | 1.9 | 3.8 | NA | NA | NA | NA | SAS, SDS | Severe |
| Chen et al., 2020b | GP | 1921 | No | NA | 0.8 | 0.0 | NA | NA | NA | NA | SAS | Severe |
| Chen et al., 2020c | GHCW | 422 | Yes | 44.6 | NA | 3.8 | NA | NA | 71.6 | NA | SDS<br>IES-R | Mild (DEP)<br>Moderate (PTSD) |
| | GP | 1071 | | | NA | 2.9 | NA | NA | 55.8 | NA | | |
| Chen et al., 2020d | FHCW | 95 | No | 74.5 | 8.5 | 8.5 | 1.1 | NA | 14.9 | NA | GAD-7, PHQ-9, ISI, PCL-C | Moderate (PTSD)<br>Severe (ANX, DEP, & INS) |
| | GHCW | 77 | | 59.7 | 0.0 | 1.3 | 0.0 | NA | 3.0 | NA | | |
| Chen et al., 2020e | GHCW | 902 | No | 68.6 | 16.6 | 18.3 | NA | NA | NA | NA | GAD-7, PHQ-9 | Moderate |
| Cheng et al., 2020 | GHCW | 534 | No | 82.4 | 0.7 | NA | 30.0 | NA | NA | NA | SAS | Moderate (INS)<br>Severe (ANX) |
| Choi et al., 2020 | GP | 500 | No | 54.8 | 14.0 | 7.0 | NA | NA | NA | NA | GAD-7, PHQ-9 | Moderate (ANX)<br>Severe (DEP) |
| Dai et al., 2020 | GHCW | 4357 | No | 76.8 | NA | NA | NA | 39.1 | NA | NA | GHQ-12 | Moderate |
| Dong et al., 2020 | FHCW | 4618 | No | 83.7 | NA | NA | NA | 24.2 | NA | NA | HEI | Moderate |
| Du et al., 2020 | FHCW | 134 | Yes | 39.4 | 20.1 | 12.7 | NA | NA | NA | NA | BAI, BDI-II | Mild |
| Elhai et al., 2020 | GP | 908 | No | 82.8 | 2.6 | 3.7 | NA | NA | NA | NA | GAD-7, DASS-21 | Severe |
| Fang, 2020 | GP | 191 | Yes | 60.2 | NA | 6.8 | NA | NA | NA | NA | PHQ-9 | Severe |
| Feng, 2020 | GP | 671 | No | 53.9 | 3.9 | 12.2 | NA | NA | 10.4 | NA | GAD-7, PHQ-2, PC-PTSD | Moderate (DEP, PTSD)<br>Severe (ANX) |
| Fong et al., 2020 | GP | 590 | No | 63.2 | NA | 15.3 | NA | NA | NA | NA | PHQ-9, | Severe |
| Fu et al., 2020 | GP | 1242 | Yes | 69.7 | 27.5 | 29.3 | 30.0 | NA | NA | NA | GAD-7, PHQ-9<br>AIS | Mild (ANX & DEP)<br>Moderate (INS) |
| Gao et al., 2020 | GP | 4872 | No | 67.7 | 22.6 | 48.3 | NA | NA | NA | NA | GAD-7, WHO-5 | Moderate |

| | | | | | | | | | | | | |
|---|---|---|---|---|---|---|---|---|---|---|---|---|
| Guo et al., 2020a | FHCW | 3351 | No | 74.8 | 2.2 | 2.5 | NA | NA | NA | NA | SAS, SDS | Severe |
| | GHCW | 7767 | | | 0.8 | 1.2 | NA | NA | NA | NA | | |
| Guo et al., 2020b | GP | 2441 | No | 52.5 | NA | 72.6 | 20.6 | NA | 79.6 | NA | CES-D-20, PSQI, PCL-5 | Moderate |
| Han et al., 2020 | FHCW | 21199 | No | 98.6 | 0.8 | 1.3 | NA | NA | NA | NA | SAS, SDS | Severe |
| Hong et al., 2020 | FHCW | 4692 | No | 96.9 | 8.1 | 9.4 | NA | NA | NA | NA | GAD-7, PHQ-9 | Moderate |
| Hou et al., 2020 | GP | 3063 | No | 56.3 | 13.3 | 14.1 | NA | NA | NA | NA | GAD-2, PHQ-2 | Moderate |
| Hu et al., 2020a | FHCW | 645 | No | 75.0 | 39.0 | 33.0 | NA | NA | NA | NA | GAD-7, PHQ-9 | Mild |
| Hu et al., 2020b | FHCW | 2014 | Yes | 87.1 | 3.3 | 1.1 | NA | NA | NA | NA | SAS, SDS | Severe |
| Huang et al., 2020a | GP | 6261 | No | 57.3 | 4.9 | 8.0 | NA | NA | NA | NA | SAS, PHQ-9 | Moderate (ANX)<br>Severe (DEP) |
| Huang et al., 2020b | FHCW | 364 | No | 58.8 | 0.8 | NA | NA | NA | NA | NA | SAS | Severe |
| Huang et al., 2020c | GP | 1172 | No | 69.3 | 33.0 | NA | 24.7 | NA | NA | NA | GAD-7, ISI | Mild (ANX)<br>Moderate (INS) |
| Jin et al., 2020 | GP | 143 | Yes | NA | 1.4 | NA | NA | NA | NA | NA | SAS | Severe |
| | | 175 | No | | 0.0 | NA | NA | NA | NA | NA | | |
| Juan et al., 2020 | FHCW | 416 | No | 70.6 | 4.6 | 6.4 | NA | NA | NA | NA | GAD-7, PHQ-9 | Moderate |
| Lai et al., 2020 | GHCW | 1257 | No | 76.7 | 5.3 | 6.2 | 1.0 | 10.5 | NA | NA | GAD-7, PHQ-9, ISI, IES-R | Severe |
| Lam et al., 2020 | FHCW | 932 | No | 75.2 | NA | 11.2 | NA | NA | NA | NA | PHQ-9 | Severe |
| Lei et al., 2020 | GP | 1593 | No | 61.3 | 0.4 | 1.1 | NA | NA | NA | NA | SAS, SDS | Severe |
| Leng et al.2020 | FHCW | 90 | Yes | 72.2 | 5.6 | NA | NA | NA | NA | NA | PCL-C | Mild |
| Li et al., 2020a | FHCW | 219 | Yes | 78.1 | NA | NA | 58.9 | NA | NA | 13.2 | SRQ-20, AIS | Moderate |
| | GHCW | 729 | No | 76.4 | NA | NA | 25.0 | 8.64 | 24.97 | 8.6 | | |
| Li et al., 2020b | GHCW | 4396 | Yes | 100.0 | 25.2 | 14.2 | NA | NA | NA | NA | GAD-7, PHQ-9 | Moderate |
| Li et al., 2020c | GP | 3637 | No | 63.0 | 16.1<br>27.5 | 22.7<br>31.2 | 26.2<br>33.7 | NA | NA | NA | GAD-7, PHQ-9<br>ISI | Mild |
| Li et al., 2020d | GP | 1109 | No | 43.9 | NA | NA | NA | NA | 67.9 | NA | IES-R | Moderate |
| Li et al., 2020e | GHCW | 908 | No | 75.6 | 0.8 | 1.2 | NA | NA | NA | NA | SAS, SDS | Severe |
| Li et al., 2020f | GP | 88611 | No | 76.9 | 13.67 | NA | NA | NA | NA | NA | GAD-7 | Moderate |
| Li et al., 2020g | FHCW | 176 | Yes | 77.3 | 50.0 | NA | NA | NA | NA | NA | HAMA | Moderate |
| Li et al., 2020h | FHCW | 225 | Yes | 72.0 | 35.6 | 46.7 | NA | NA | 31.6 | NA | DASS-21, IES-R | Mild (ANX & DEP)<br>Moderate (PTSD) |

| | | | | | | | | | | | | |
|---|---|---|---|---|---|---|---|---|---|---|---|---|
| Li et al., 2020i | FHCW | 356 | No | 86.2 | NA | NA | NA | NA | 61.8 | NA | PCL-5 | Moderate |
| Li et al., 2020j | FHCW | 150 | No | 62.7 | 39.3 | 44.7 | NA | NA | NA | NA | HAMA, HAMD | Moderate |
| Liang et al., 2020 | FHCW | 168 | No | 79.2 | 19.6 | 29.2 | 13.1 | NA | NA | NA | GAD-7, PHQ-9, ISI | Moderate |
| | GP | 1835 | | 74.4 | 10.8 | 23.1 | 8.6 | NA | 8.61 | NA | | |
| Lin et al., 2020a | GHCW | 2316 | No | NA | 41.1 | 46.9 | 32.0 | NA | NA | NA | GAD-7, PHQ-9, ISI | Mild |
| Lin et al., 2020b | GP | 5461 | No | 70.1 | 9.2 | 10.8 | 2.7, 1.6 | NA | NA | NA | GAD-7, PHQ-9, ISI | Severe |
| Liu et al., 2020a | GHCW | 512 | No | 84.6 | 0.8 | NA | NA | NA | NA | NA | SAS | Severe |
| Liu et al., 2020b | GP | 14592 | No | 68.4 | 6.6 | 10.1 | NA | NA | NA | 3.2 | GAD-7, PHQ-9, SRQ-20 | Severe |
| Liu et al., 2020c | GP | 285 | Yes | 54.4 | NA | NA | NA | NA | 7.0 | NA | PCL-5 | Moderate |
| Liu et al., 2020d | GHCW | 4976 | No | 82.3 | 2.2 | 2.3 | NA | NA | NA | 15.9 | SAS, SDS, SRQ-20 | Moderate (GPS)<br>Severe (ANX & DEP) |
| Liu et al., 2020e | GP | 4991 | No | 50.4 | 1.50 | NA | NA | NA | NA | NA | SAS | Severe |
| Liu et al., 2020f | GP | 574 | No | 57.7 | NA | 0.7 | NA | NA | NA | NA | SDS | Severe |
| Liu et al., 2020g | FHCW | 317 | No | 80.2 | 12.7 | 18.9 | NA | NA | NA | NA | GAD-7, PHQ-9 | Moderate |
| | GHCW | 719 | | | 13.6 | 18.2 | NA | NA | NA | NA | | |
| Liu et al., 2020h | FHCW | 742 | No | 85.5 | 4.0 | 2.1 | NA | NA | NA | NA | DASS-21 | Severe |
| | GHCW | 1289 | | | 1.2 | 1.0 | NA | NA | NA | NA | | |
| Lu et al., 2020a | GHCW | 2042 | No | 77.6 | 2.9 | 0.3 | NA | NA | NA | NA | HAMA, HAMD | moderate (ANX)<br>severe (DEP) |
| | GP | 257 | | | 1.6 | 0.0 | NA | NA | NA | NA | | |
| Lu et al., 2020b | GHCW | 1848 | No | 63.0 | NA | 18.8 | NA | NA | NA | NA | CES-D-9 | Moderate |
| Lu et al., 2020c | FHCW | 382 | Yes | 61.8 | 7.6 | 9.9 | NA | NA | 6.8 | NA | GAD-7, PHQ-9, PCL-C | Moderate (PTSD)<br>Severe (ANX and DEP) |
| | GP | 1035 | | 91.4 | 6.3 | 6.8 | NA | NA | 4.5 | 4.50 | | |
| Mi et al., 2020 | GHCW | 1029 | No | 61.6 | 6.6 | 13.3 | NA | NA | NA | NA | GAD-2, HQ-2 | Moderate |
| Ni et al., 2020a | GHCW | 214 | Yes | 60.8 | 23.8 | 19.2 | NA | NA | NA | NA | GAD-2, PHQ-2 | Moderate |
| | GP | 1577 | | 68.8 | 22.0 | 19.2 | NA | NA | NA | NA | | |
| Ni et al., 2020b | GP | 2551 | No | 68.9 | 1.8 | 14.9 | NA | NA | NA | NA | GAD-7, PHQ-2 | Moderate (DEP)<br>Severe (ANX) |
| Ning et al., 2020 | GHCW | 612 | No | 72.9 | 16.3 | 25.0 | NA | NA | NA | NA | SAS, SDS | Mild |
| Pan et al., 2020a | GHCW | 194 | Yes | 81.8 | 3.6 | 5.2 | NA | NA | NA | NA | GAD-7,PHQ-9 | Moderate |
| Pan et al., 2020b | GP | 3035 | No | 46.9 | NA | 5.6 | NA | NA | NA | NA | PHQ-9 | Moderate |

| | | | | | | | | | | | | |
|---|---|---|---|---|---|---|---|---|---|---|---|---|
| Qi et al., 2020 | FHCW | 801 | No | 79.9 | NA | NA | 78.4 | NA | NA | NA | PSQI | Moderate |
| | GHCW | 505 | | 81.2 | NA | NA | 61.0 | NA | 61.00 | NA | | |
| Qian et al., 2020a | GP | 510 | Yes | 50.0 | 32.7 | NA | NA | NA | NA | NA | GAD-7 | Moderate |
| | | 501 | No | 49.1 | 20.4 | NA | NA | NA | NA | NA | | |
| Qian et al., 2020b | GP | 510 | Yes | 50.0 | 32.8 | NA | NA | NA | NA | NA | GAD-7 | Moderate |
| | | 501 | No | 49.1 | 20.5 | NA | NA | NA | NA | NA | | |
| Qiu et al., 2020 | GP | 52730 | No | 64.7 | NA | NA | NA | 5.14 | NA | NA | CPDI | Severe |
| Que et al et al., 2020 | GHCW | 2285 | No | 69.1 | 11.6 | 12.8 | 6.8 | NA | NA | NA | GAD-7, PHQ-9, ISI | Moderate |
| Ren et al., 2020 | GP | 6130 | No | 66.9 | 7.1 | 12.0 | NA | NA | NA | NA | GAD-7, PHQ-9 | Moderate |
| Shi et al., 2020 | FHCW | 9725 | No | 52.1 | 11.9 | 12.8 | 7.0 | NA | NA | NA | GAD-7, PHQ-9 | Moderate |
| | GP | 46954 | | | 10.0 | 10.4 | 5.5 | NA | 5.49 | NA | | |
| Si et al., 2020 | GHCW | 863 | No | 70.7 | 13.6 | 13.9 | NA | NA | 40.2 | NA | DASS-21, IES-6 | Mild (ANX & DEP)<br>Moderate (PTSD) |
| Song et al., 2020a | GHCW | 14825 | No | 64.3 | NA | 25.2 | NA | NA | 9.1 | NA | PCL-5, CES-D-20 | Moderate |
| Song et al., 2020b | GP | 709 | No | 74.2 | 12.7 | 13.5 | 20.7 | NA | NA | NA | GAD-7, CES-D-10, ISI | Mild (INS), Moderate (ANX & DEP) |
| Su et al., 2020 | GP | 403 | No | 68.5 | 5.0 | NA | NA | NA | NA | NA | GAD-7 | Severe |
| Sun et al., 2020a | GP | 2091 | No | 60.8 | NA | NA | NA | NA | 4.6 | NA | PCL-5 | Moderate |
| Sun et al., 2020b | FHCW | 170 | Yes | 92.4 | 1.2 | 0.0 | NA | NA | NA | NA | DASS-21 | Severe |
| Sun et al., 2020c | GP | 472 | No | 56.4 | NA | 75.4 | NA | NA | NA | NA | CES-D-10 | Mild |
| Sun et al., 2020d | GHCW | 536 | No | 69.0 | 9.7 | 18.8 | NA | NA | NA | NA | GAD-7, PHQ-9 | Mild |
| Tan et al., 2020 | GP | 673 | No | 25.6 | 1.3 | 0.9 | 0.4 | NA | 10.8 | NA | DASS-21, ISI, IES-R | Moderate (PTSD), Severe (ANX, DEP, & INS) |
| Teng et al., 2020 | GHCW | 398 | No | 75.9 | 0.3 | 4.3 | NA | NA | NA | NA | SAS, PHQ-9 | Severe |
| Tu et al., 2020 | GHCW | 100 | No | 100.0 | 10.0 | 2.0 | 2.0 | NA | NA | NA | GAD-7, PHQ-9, PSQI | Severe |
| Wang et al., 2020a | GP | 600 | No | 55.5 | 0.0 | 0.3 | NA | NA | NA | NA | SAS, SDS | Severe |
| Wang et al., 2020b | GHCW | 123 | Yes | 90.0 | 7.3 | 25.2 | 38.2 | NA | NA | NA | SAS, SDS, PSQI | Mild (ANX & DEP)<br>Moderate (INS) |
| Wang et al., 2020c | GHCW | 1045 | No | 85.8 | 20.0 | 13.6 | 10.4 | NA | NA | NA | HADS, ISI | Moderate |
| Wang et al., 2020d | GP | 6437 | No | 56.1 | NA | NA | 17.7 | NA | NA | NA | PSQI | Moderate |
| Wang et al., 2020e | FHCW | 274 | No | 77.4 | NA | NA | 19.7 | NA | NA | NA | PSQI | Moderate |

| | | | | | | | | | | | | |
|---|---|---|---|---|---|---|---|---|---|---|---|---|
| Wang et al., 2020f | FHCW | 179 | No | 52.0 | 14.5 | 5.3 | 7.3 | NA | NA | NA | GAD-7, PHQ-9<br>ISI | Severe |
| | GP | 16066 | | | 2.7 | 2.0 | 3.1 | NA | NA | NA | | |
| Wang et al., 2020g | FHCW | 661 | No | 64.5 | 24.8 | 36.6 | 23.6 | NA | NA | NA | HADS, PSQI | Mild (ANX & DEP)<br>Moderate (INS) |
| | GHCW | 853 | | | 22.3 | 36.6 | 17.0 | NA | NA | NA | | |
| | GP | 487 | | | 20.1 | 31.8 | 13.3 | NA | NA | NA | | |
| Wang et al., 2020h | FHCW | 202 | No | 87.6 | NA | NA | NA | NA | 9.4 | NA | PCL-C | Moderate |
| Wang et al., 2020i | GP | 2540<br>2543 | No | NA | 32.0<br>36.4 | 36.1<br>42.9 | NA | NA | NA | NA | GAD-7, PHQ-9 | Mild |
| Wang et al., 2020j | FHCW | 742 | Yes | 82.5 | 38.5 | 21.7 | 15.4 | NA | NA | NA | GAD-7, PHQ-9, ISI | Mild (ANX & PTSD)<br>Moderate (DEP) |
| | GHCW | 1155 | | | 19.8 | 10.7 | 6.1 | NA | NA | NA | | |
| Wu et al., 2020a | GP | 24789 | No | 46.3 | 3.6 | 3.1 | NA | NA | NA | NA | HADS | Severe |
| Wu et al., 2020b | GHCW | 304 | No | 68.1 | NA | NA | NA | 14.1 | NA | NA | K-6 | Moderate |
| Wu et al., 2020c | GP | 104 | Yes | 57.7 | NA | NA | NA | NA | 9.4 | NA | PCL-5 | Moderate |
| | | 330 | No | 67.3 | NA | NA | NA | NA | 7.2 | 7.20 | | |
| Xiao et al., 2020 | GHCW | 958 | No | 67.2 | 54.1 | 57.3 | NA | NA | NA | NA | HADS | Mild |
| Xu et al., 2020 | GHCW | 8817 | No | 78.0 | 0.0 | 3.2 | NA | NA | NA | NA | GAD-7, PHQ-9 | Severe |
| Xing et al., 2020 | FHCW | 309 | No | 97.4 | 2.3 | 0.6 | NA | NA | NA | NA | SAS, SDS | Severe |
| Xiong et al., 2020 | GHCW | 231 | No | 97.3 | 3.6 | 1.8 | NA | NA | NA | NA | GAD-7, PHQ-9 | Severe |
| Yang et al., 2020a | GHCW | 449 | No | NA | 29.2 | NA | NA | NA | NA | NA | SAS | Mild |
| Yang et al., 2020b | GP | 2410 | No | 49.2 | NA | NA | 14.9 | NA | NA | NA | PSQI | Mild |
| Yin et al., 2020a | GP | 8151 | No | 57.7 | 6.7 | 13.5 | NA | NA | NA | NA | GAD-7, PHQ-9 | Moderate |
| Yin et al., 2020b | GHCW | 371 | No | 61.5 | NA | NA | NA | NA | 3.8 | NA | PCL-5 | Moderate |
| Ying et al., 2020 | GP | 845 | No | 47.3 | 33.7 | 29.4 | NA | NA | NA | NA | GAD-7, PHQ-9 | Mild |
| Yu et al., 2020a | GP | 1588 | No | 66.9 | NA | NA | NA | 22.8 | NA | NA | K-6 | Moderate |
| Yu et al., 2020b | GP | 1138 | No | 65.6 | NA | NA | 29.9 | NA | NA | NA | ISI | Moderate |
| Yu et al., 2020c | FHCW | 290 | No | 64.1 | 7.3 | NA | NA | NA | NA | NA | GAD-7 | Severe |
| Zhan et al., 2020a | FHCW | 2667 | Yes | 97.0 | 39.8 | 54.7 | NA | NA | NA | NA | GAD-7, PHQ-9 | Moderate |
| Zhan et al., 2020b | FHCW | 1794 | Yes | 97.0 | NA | NA | 52.8 | NA | NA | NA | AIS | Moderate |
| Zhang et al., 2020a | GHCW | 1563 | No | 82.5 | 44.7 | 50.7 | 36.1 | 73.4 | NA | NA | GAD-7, PHQ-9, ISI, IES-R | Mild |
| Zhang et al., 2020b | GP | 98 | No | 65.3 | 16.3 | 14.3 | NA | NA | NA | NA | GAD-7, PHQ-9 | Severe |

| | | | | | | | | | | | | |
|---|---|---|---|---|---|---|---|---|---|---|---|---|
| Zhang et al., 2020c | GP | 369 | No | 45.0 | NA | NA | NA | 0.0 | NA | NA | K-6 | Severe |
| Zhang et al., 2020d | GHCW | 927 | No | 73.1 | 13.0 | 12.2 | 38.4 | NA | NA | NA | GAD-2, PHQ-2, ISI | Mild (INS)<br>Moderate (ANX & DEP) |
| | GP | 1255 | | 57.6 | 8.5 | 9.5 | 30.5 | NA | NA | NA | | |
| Zhang et al., 2020e | GP | 263 | No | 59.7 | NA | NA | NA | NA | 7.6 | NA | IES-R | Moderate |
| Zhang et al., 2020f | FHCW | 966 | No | 76.4 | 10.7 | 17.3 | NA | NA | NA | NA | GAD-7, PHQ-9 | Moderate |
| Zhang et al., 2020g | FHCW | 421 | Yes | 85.1 | 14.6 | 16.2 | 2.8 | NA | 22.6 | NA | HADS, ISI<br>PCL-C | Moderate (ANX, DEP, & PTSD), Severe (INS) |
| | GP | 221 | | | | | NA | NA | 17.7 | NA | | |
| Zhang et al., 2020h | GP | 1342 | No | 62.7 | NA | 13.60 | NA | NA | NA | NA | PHQ-9 | Moderate |
| Zhang et al., 2020i | GP | 123768 | No | 29.4 | 0.3 | 0.1 | NA | NA | NA | NA | SAS, SDS | Severe |
| Zhang et al., 2020j | FHCW | 269 | No | 47.1 | 5.6 | 4.8 | 7.8 | NA | NA | NA | GAD-7, PHQ-9, ISI | Severe |
| | GP | 2640 | | | 4.4 | 3.1 | 5.6 | NA | NA | NA | | |
| Zhao et al., 2020a | FHCW | 972 | No | 62.7 | 16.9 | 10.3 | 11.2 | NA | NA | NA | GAD-7, PHQ-9, ISI | Moderate |
| Zhao et al., 2020b | GP | 1501 | No | NA | 15.8 | 14.8 | NA | NA | NA | NA | GAD-2, PHQ-2 | Moderate |
| Zhao et al., 2020c | GP | 1630 | No | NA | 0.8 | NA | 36.4 | NA | NA | NA | SAS, PSQI | Mild, Severe |
| Zhou et al., 2020a | FHCW | 1931 | No | 12.0 | NA | NA | 18.4 | NA | NA | NA | PSQI | Moderate |
| Zhou et al., 2020b | FHCW | 606 | No | 81.2 | 45.4 | 57.6 | 32.0 | NA | NA | NA | GAD-7, PHQ-9, ISI | Mild |
| | GP | 1099 | | 69.4 | 33.8 | 47.6 | 25.1 | NA | NA | NA | | |
| Zhu et al., 2020a | GP | 5281 | No | 85.0 | 24.1 | 13.5 | NA | NA | NA | NA | GAD-7, PHQ-9 | Moderate |
| Zhu et al., 2020b | GHCW | 453 | No | 94.9 | 28.6 | NA | NA | NA | NA | NA | SAS | Moderate |
| Zhu et al., 2020c | FHCW | 165 | No | 83.0 | 11.4 | 45.6 | NA | NA | NA | NA | SAS, SDS | Mild |
| Zhu et al., 2020d | FHCW | 320 | No | 63.7 | 19.7 | 21.3 | NA | NA | NA | 12.8 | GAD-7, PHQ-9, SRQ-20 | Mild (ANX & DEP<br>Moderate (GPS) |
| | GHCW | 538 | | 73.0 | 17.1 | 18.0 | NA | NA | NA | 10.0 | | |

Note: ANX= Anxiety, FHCW=Frontline Healthcare Worker, GHCW = General Healthcare Worker, GP = General Population, INS= INS, GPS=General psychological symptoms, DEP= Depression, DIS=DIS, NA= Not available, Mild = Above Mild, Moderate = Above Moderate, Severe =Above severe

**Table S3. The instruments of mental health disorders in the individual papers in the systematic review**

| Instrument | Frequency | Percent | Citation |
|---|---|---|---|
| **Anxiety** | 127 | 100 | |
| PHQ (GAD-7/GAD-2, Generalized Anxiety Disorder scale - 7-item/2-item) | 77 | 60.6 | 157 158 |
| SAS (Self-rating Anxiety Scale) | 30 | 23.6 | 159 |
| HADS (Hospital Anxiety Depression Scale) | 7 | 5.5 | 160 |
| DASS-21 (Depression Anxiety Stress Scale) | 6 | 4.7 | 161 |
| HAMA (Hamilton Anxiety Rating Scale) | 6 | 4.7 | 162 |
| BAI (Beck Anxiety Inventory) | 1 | 0.8 | 163 |
| **Depression** | 128 | 100 | |
| PHQ (PHQ-9/PHQ-2, The Patient Health Questionnaire depression scale - 9-item/2-item) | 81 | 63.3 | 158 164 |
| SDS (Self-rating Depression Scale) | 17 | 13.3 | 165 166 |
| CES-D (CES-D-20/9/10, The Center for Epidemiologic Studies Depression Scale – 20 items/9 items/10 items) | 9 | 7.0 | 167 |
| DASS-21 (Depression Anxiety Stress Scale) | 7 | 5.5 | 161 |
| HADS (Hospital Anxiety Depression Scale) | 7 | 5.5 | 160 |
| HAMD (Hamilton Depression Scale) | 5 | 3.9 | 168 |
| BDI-II | 1 | 0.8 | 169 |
| WHO-5 | 1 | 0.8 | 170 |
| **Distress** | 9 | 100 | |
| K-6 (the six-item Kessler mental distress scale) | 4 | 44.4 | 171 |
| IES-R (Impact of Event Scale - Revised) | 2 | 22.2 | 172 |
| CPDI (COVID-19 Peritraumatic Distress Index) | 1 | 11.1 | 50 |
| GHQ-12 (General Health Questionnaire) | 1 | 11.1 | 22 |
| HEI (Huaxi Emotional-Distress Index) | 1 | 11.1 | 173 |
| **General psychological symptoms** | | | |
| SRQ-20 (Self Reporting Questionnaire 20) | 7 | 100 | 174 |
| **Insomnia** | 57 | 100 | |
| ISI (Insomnia Severity Index) | 36 | 63.2 | 175 |
| PSQI (Pittsburgh Sleep Quality Index) | 17 | 29.8 | 176 177 |
| AIS (Athens Insomnia Scale) | 4 | 7.0 | 178 |
| **PTSD** | 30 | 100 | |
| IES-R (Impact of Event Scale – Revised) | 12 | 40.0 | 179-181 |
| PCL-5 (the Posttraumatic Stress Disorder Checklist for DSM-5) | 8 | 26.7 | 182 |
| PCL-C (Posttraumatic Stress Disorder Checklist-Civilian Version) | 8 | 26.7 | 183-185 |
| IES-6 (Impact of Event Scale – 6 items) | 1 | 3.3 | 186 |
| PC-PTSD (The Primary Care PTSD Screen for DSM-5) | 1 | 3.3 | 187 |
